# Assessing Clinical Progression Measures in Alzheimer’s Disease Trials: A Systematic Review and Meta-Analysis

**DOI:** 10.1101/2023.08.29.23294771

**Authors:** Jonathan McLaughlin, William J. Scotton, John A. Hardy, Maryam Shoai

**Affiliations:** NHS Research Scotland (NRS) Career Research Fellow, Department of Psychological Medicine, Aberdeen Royal Infirmary, Aberdeen, UK; Department of Neurodegenerative Diseases, UCL Queen Square Institute of Neurology, University College London, London, UK; UK Dementia Research Institute, University College London, London, UK; Aligning Science Across Parkinson’s (ASAP) Collaborative Research Network, Chevy Chase, MD, 20815; Reta Lila Weston Institute, UCL Queen Square Institute of Neurology, London, UK; National Institute for Health Research (NIHR) University College London Hospitals Biomedical Research Centre, London, UK; Institute for Advanced Study, The Hong Kong University of Science and Technology, Hong Kong SAR, China

## Abstract

Clinical trials in Alzheimer’s disease (AD) aim to reduce the rate of progression of disease. This is heavily dependent on a consensus of a minimum clinically important difference as well as the ability of the cognitive and functional measures used to accurately measure progression.

In this study we perform a systematic review and meta-regression to assess the precision of measurement of AD clinical progression in clinical trials of therapeutic interventions in patients with known positive amyloid status prior to trial entry.

Meta analyses of randomised controlled trials (RCT) in AD, with amyloid positive status (Aβ+) as an inclusion criterion, were undertaken with functional, cognitive, and composite measures included in the analyses. Twenty-five RCTs were eligible for inclusion. Whilst most RCTS enrolled prodromal or mild AD patients with an average MMSE score at baseline of 27, several included average MMSE scores as low as 22. We performed meta regressions, correcting for age, gender, and stage of disease in R version 4.2.0, using the *metafor* and *emmeans* libraries. Of the progression measures included in the meta-analyses, the FAQ, a functional measure, had the largest weighted mean change over 12-weeks followed by MMSE, whilst the most commonly used neuropsychiatric battery, NPI, failed to show sensitivity to change in the given time period. This study emphasises the necessity of appropriate composite progression measures that weigh cognitive, functional and neuropsychiatric symptoms according to their ability to detect meaningful change in symptoms and thus have a better chance of detecting meaningful change in participants of interventional RCTs.

**Summary:** *Background:* Alzheimer’s disease (AD) is a slowly progressive disease. It is now widely recognised that there is a pre-clinical phase. This phase of the disease may be apparent via biomarker testing up to 20 years before clinically evident AD. Pre-clinical AD is then followed by clinically significant cognitive decline ranging from MCI to severe AD. The aim of randomised controlled trials (RCT) is to reduce or halt the rate of clinical progression of AD. Most of these trials have been unsuccessful. To determine the effectiveness of treatments there must be robust and reliable tools for measuring AD progression. For at least 30 years there has been recognition that the measures of progression used in AD clinical trials are problematic. A significant concern is that current measures of clinical progression are potentially not sensitive enough in early and preclinical stages of AD and so are not reliable indicators of AD progression. In this systematic review and meta-regression we aimed to assess the precision of measurements of clinical progression in AD clinical trials of therapeutic interventions in patients with known positive amyloid status prior to trial entry.

*Methods:* Meta analyses of RCTs in AD with amyloid positive status (Aβ+) as an inclusion criterion was undertaken with functional, cognitive, and composite measures included in the analyses. Twenty-five RCTs were eligible for inclusion. Statistical analyses were performed using R version 4.2.0 and the *metafor* and *emmeans* libraries.

*Findings:* Of the progression measures commonly reported in RCTs, the FAQ, had the largest weighted mean change over 12-weeks followed by MMSE. Other cognitive measures were amongst the least sensitive measures over the chosen time period. As a composite score, both the iADRS and CDRSB appear to be performing better than the cognitive components they comprise. The neuropsychiatric battery analysed in this study appeared to be the least sensitive of measures of progression.

*Interpretation:* Functional measures, with the exception of QoL-AD, perform better than other groups of measures. Measures which rely on purely cognitive domains are not optimal for sole use in AD trials. Ideally, measures should include both cognitive and functional components to enhance sensitivity. New composite measures address the poorer performance of composite scores, as compared to their comprising functional measures, by assigning different weights to cognitive and functional change.

## Introduction

As treatments for AD were sought and developed there was a need to have instruments which could accurately measure clinical progression in AD. Some measures, which had been initially conceived as clinical screening tools, served this purpose, *e.g.* Mini Mental State Examination (MMSE). Other measures were specifically conceived to measure AD progression. The abbreviations for each measure are listed in *Appendix 1*.

The aim of clinical trials is to reduce or halt the rate of clinical progression of AD. Many of these trials have been unsuccessful. To determine the effectiveness of treatments there must be robust and reliable tools for measuring AD progression. For at least 30 years there has been recognition that the measures of progression used in AD clinical trials are problematic [1]. One criticism is that clinical measures of progression are affected by symptomatic treatments (*e.g.*, Cholinesterase Inhibitors (ChEIs)) and therefore the true disease modifying effects of potential therapies are masked [2]. Another, more significant concern, is that the most commonly used measures of clinical progression are potentially not sensitive enough in early and preclinical stages of AD and so are not reliable indicators of AD progression [3].

These criticisms have led to the development of newer composite measures as endpoints in clinical trials [4]. One of the issues though is the lack of a standard adopted composite score in AD clinical trials and therein the proliferation of many different composite measures of AD progression. This makes comparison between studies very difficult.

This has led to a focus on biomarkers which can act as surrogate outcome markers in therapeutic trials. This shift in focus has been seen most clearly in the case of anti-amyloid therapies which act to reduce amyloid load in the brain. However, as highlighted by the controversy around the clinical utility of these therapeutic agents [5], it is not clear that amyloid reduction, or clinical rating scale change is linked to meaningful clinical improvement [6]. Indeed, clinical trial participants, in whom amyloid plaque reduction has been demonstrated still experience progressive neurodegeneration [7]. Much of the difficulty in trying to develop measures of progression in AD stems from the clinical and pathological heterogeneity of the disease. There are different sources of this, including; genetics, neuropathology and demographics [8]. There has been a subsequent drive to enrich trial designs to account for these factors.

## Rationale and aims of the review

It is not known quantitatively if some clinical progression measures perform better than others in AD clinical trials. This performance rests on the sensitivity of a measure to detect change over time and that this change is clinically meaningful. By analysing the placebo groups of clinical trials, which have confirmed Aβ pathology in their included participants, this review aims to compare sensitivity of progression measures at detecting change and to suggest if particular progression measures are better placed than others to be utilised in AD clinical trials.

## Methods

### Search strategy and selection criteria

A search of the International Prospective Register of Systematic Reviews (PROSPERO) and The Cochrane Library revealed that no similar review had been undertaken.

The research question was formulated ahead of the search strategy. The Population, Intervention, Control, Outcomes and Study Design principle (PICOS) informed this:

#### Population

Males and Females over 18 years of age with a diagnosis of AD, MCI due to AD pathology, Prodromal AD or Preclinical AD in whom AD pathology has been established by the presence of elevated levels of amyloid protein on Amyloid PET or via CSF analysis prior to inclusion in a clinical trial. Only placebo groups were used in the review.

#### Intervention

Studies investigating therapeutic agents for the treatment of Alzheimer’s disease whether this be in established AD or Prodromal AD or MCI due to AD pathology.

#### Control

Not applicable – the placebo groups in the therapeutic trials formed the target population for this review.

#### Outcomes

The assessment of progression in Alzheimer’s disease via novel clinical progression measures/models or established clinical measures (*e.g.* ADAS-COG, MMSE, CDR-SB).

#### Study design

Randomised Controlled Trials (RCTs) were exclusively sought. This process refined the research question underpinning the systematic review; *To analyse the methods of measuring clinical progression in Randomised Controlled Trials in Alzheimer’s Disease, specifically using placebo arm patients with known positive amyloid status prior to trial entry*.

### Inclusion and Exclusion Criteria

Studies were included if they met the defined criteria as follows:

- RCT in design and trialling a therapeutic intervention.
- RCTs that used clinical outcome/progression measures.
- Participant inclusion criteria stipulated that only Aβ+ subjects/subjects with elevated Aβ were included in the trial as determined by either Amyloid PET or CSF analysis.
- Subjects had a diagnosis of AD, MCI due to AD pathology, Prodromal AD or were Cognitively Normal (CN) but with evidence of elevated Aβ (Preclinical AD).
- The RCT must have a placebo group and have reported data on clinical outcome/progression measures for the placebo group. In circumstances where the RCT did not have a published report in a peer reviewed journal it could be included if data was reported on a recognised forum such as ClinicalTrials.gov.

Studies were excluded if they met the defined criteria as follows:

- Observational, case study or reports, case control or cohort study in design.
- RCTs which report elevated tau levels but in which there is no account of Aβ*+*.
- RCTs which included patients with other forms of dementia/pathology *e.g.* suspected cerebral vascular disease.
- RCTs which trialled an intervention for symptoms/difficulties arising other than cognitive decline.
- RCTs which trialled an intervention aimed at caregivers *e.g.,* education intervention.
- Insufficient data reported *e.g.,* no data on change in clinical measures from baseline to endpoint or idiosyncratic and sparse clinical measures used.

No limits were placed on the number of trial participants, purported severity of AD, length of the trial, class of therapeutic agent, the main outcome of the trial, and the number of clinical outcome/progression measures used.

### Resources for search

The following resources were used for the study search; MEDLINE (Ovid interface), Embase (OVID interface), PubMed, Google Scholar, National Institute of Health ClinicalTrials.gov, AIBL list of publications, International Clinical Trials Registry Platform (ICTRP WHO), International Standard Randomised Controlled Trial Number registry (ISRCTN), European Union Drug Regulating Authorities Clinical Trials Database (EudraCT) and Researchregistry.com.

### Scoping search

An initial scoping electronic search was devised and refined, *Appendix 2* The purpose was to define and optimise search terms, ensure no similar and previous review had been undertaken (prior PROSPERO and Cochrane library searches had suggested not) and to establish whether there was enough literature to proceed. On this later point, and on review of titles and abstracts of the 2,744 records found in the scoping search, 117 studies were identified for full text review. Of these only 3 records fulfilled criteria for inclusion. The scoping search was successful in helping to identify relevant keywords and Medical Subject Headings (MESH terms) as well as allowing identification of related reviews which had published their search strategies e.g. [2]. Additionally, via scoping it became clear that a significant proportion of trials that met the inclusion criteria would potentially be subject to publication bias as they had statistically non-significant outcomes. This, in part, would explain the low rate of studies found on database searching.

### Search Protocol

“Preferred Reporting Items for Systematic Reviews and Meta-Analyses” (PRISMA) [9] were utilised to develop the methodology and reporting.

***Figure 1*** provides a visual summary of the procedure in selecting studies for inclusion in the systematic review.

**Figure 1:**
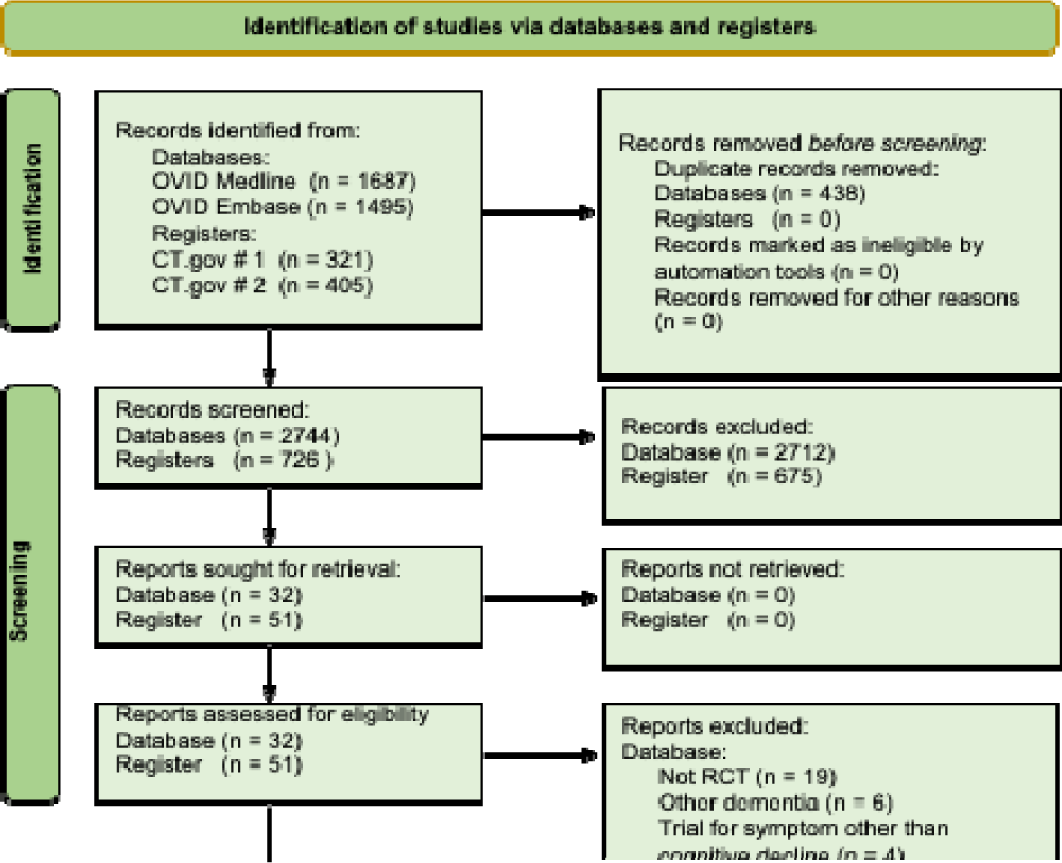
PRISMA flow diagram.

### Risk of Bias

Included studies were assessed for bias in line with Cochrane guidelines for the reporting and conduct of systematic reviews, using the Cochrane Risk of Bias tool for randomised trials version 2 (RoB 2) [10]. This analysis revealed some potential bias arising from deviations from the intended interventions as well as bias in selection of reported results. However, other domains such as bias arising from the randomisation process as well as bias due to missing data performed strongly and therefore, we felt the results of the studies could be utilised in the analysis.

### Data extraction process

Data was manually extracted from either the published study or from the data published in the registry entry on ClinicalTrials.gov. The data extracted pertained to the placebo group participants only. Data was recorded in a Microsoft excel spreadsheet for each reported outcome measure with 2 or more studies. Baseline characteristics were also recorded for the placebo group subjects. Outcome measures were reported in 2 principal ways. The commonest method was to report arithmetic mean (AM) change in an outcome measure from baseline to a time endpoint with either Standard Error (SE), Standard Deviation (SD) or 95% Confidence Intervals (95% CI). 13 of the 25 included trials used this method. The other method, used in 11 trials, was the reporting of change in outcome measures over time as the Least Squares Mean (LSM) with either SE, SD or 95% CI. One study reported mean raw outcome measure scores at baseline and at endpoint with SD [11]. From the 25 included studies there were 26 data sets for analysis. A possible 27^th^ data set was requested from a corresponding author [12] for the Margeurite Road trial but no response was received.

### Statistical Analysis

Statistical analyses were performed in R version 4.2.0 [13] using the *metafor, and emmeans* package [14, 15] which was updated to include inverse variance in order to allow for inclusion of heterogenous studies in meta-analyses. Outcome measure change was reported as either an adjusted mean, LSM, or arithmetic mean, or as mean change in outcome score. This data was extracted from the corresponding published report or published completed trial results where a report was not available.

As well as being a reasonable timeframe over which progression effects may be observable; a period of 12 weeks was chosen to maximise available trial data. Mean change in the outcome measure per 12 weeks was calculated for the data obtained.

Due to the expected high heterogeneity of outcome measures, i.e. LSM vs AM, we opted for a random effects model with an inverse variance heterogenous meta-analysis known as IVhet [16]. Heterogeneity of variance across included studies was assessed using the I^2^ and Q-statistics. Influential studies and small study bias was assessed.

Moderator variables were also assessed and included the mean age of placebo participants at baseline, the percentage of females constituting the placebo groups, and AD stage targeted as ascertained by mean MMSE score at baseline. It was felt that all three factors could reasonably be expected to influence the weighted mean change in outcome measures. Thus, meta regressions were performed with percentage female, mean age at baseline, and AD stage when variation existed. The *emmeans* package was used to predict the estimated value of mean change under a model that assumes mean of continuous moderator and the mean of the dummy variables for each level of the categorical moderator.

## Results

Twenty-five RCTs and twelve measures were included. The included trials span a publication period from 2015 to 2022. Fifteen trials had trial registration, published trial data on ClinicalTrials.gov and a peer reviewed linked publication in a journal. The characteristics of the placebo groups are summarised in ***Table 1***.

**Table 1:** Summary of baseline characteristics of placebo groups in the included studies.

| Study name | Placebo<br>N | Females<br>% | Age (SD) | MMSE | Diagnosis |
| --- | --- | --- | --- | --- | --- |
| Mullins<br>2019 | 14 | 60% | 74.0 (6.4) | > 20 | MCI or early AD |
| Eli Lilly<br>2018 | 20 | 55% | 67.73 (7.1) | 20-26 Probable<br>AD 27-30 MCI<br>due to AD | MCI due to AD or<br>Probable AD<br>NINCDS/ADRDA |
| ENGAGE<br>2021 | 545 | 52.70% | 69.8 (7.7) | 24-30 inclusive | MCI due to AD or mild<br>AD NIAAA |
| EMERGE<br>2021 | 548 | 52.90% | 70.8 (7.4) | 24-30 inclusive | MCI due to AD or mild<br>AD NIAAA |
| Sperling<br>2021 | 185 | 58.40% | 70.2 (5.8) | NA | Clinically normal |
| CREAD<br>2020 | 409 | 60.40% | 70.3 (8.4) | ≥22 | Probable or Prodromal<br>AD NIAAA |
| NAVIGATE-AD<br>2021 | 133 | 58 | 72.54 (7.8) | 20-26 inclusive | Mild AD NIAAA |
| CREAD 2<br>2020 | 399 | 56.4 | 70.7 (7.9) | ≥ 22 | Probable or Prodromal<br>AD NIAAA |
| **Novartis**<br>**2021** | 456 | 63.2 | 65-70 | $\geq 24$ | Cognitively unimpaired |
| **Teng**<br>**2022** | 135 | 55.6 | 69.7 (7.3) | $\geq 20$ | Probable or Prodromal<br>AD NIAAA |
| **MissionAD 1 & 2**<br>**2021** | 1108 | 53.6 | 72.1 (7.1) | $\geq 24$ | MCI due to AD or mild<br>AD NIAAA |
| **AMARANTH**<br>**2019** | 740 | 53.8 | 71.4 (6.9) | 20-30 inclusive | Probable AD or MCI<br>due to AD NIAAA |
| **DAYBREAK**<br>**2019** | 562 | 61.9 | 72.1 (7.1) | 20-26 inclusive | Probable AD or MCI<br>due to AD NIAAA |
| **APECS**<br>**2019** | 485 | 43.9 | 71.6 (7.1) | $\geq 24$ | "Prodromal AD" |
| **Coric**<br>**2015** | 131 | 42.0 | 71.6 (7.8) | 24-30 inclusive | "Prodromal AD" |
| **EXPEDITION 3**<br>**2018** | 1072 | 58.9 | 73.26 (8.0) | 20-26 inclusive | Probable AD<br>NINCDS/ADRDA |
| **Swanson**<br>**2021** | 238 | 59.0 | 71 (NA) | 22-30 inclusive | MCI due to AD or Mild<br>AD NIAAA |
| **Potter**<br>**2021** | 20 | 55.0 | 70.15 (6.4) | 10-26 inclusive | Moderate AD MMSE<br>criteria |
| **van Dyck**<br>**2016** | 21 | 42.9 | 69.6 (6.8) | $\geq 25$ | Subjective memory<br>complaint & no<br>dementia diagnosis |
| **SCarlet RoAD**<br>**2021** | 266 | 56 | 69.5 (7.5) | $\geq 24$ | Prodromal AD<br>NINCDS/ADRDA |
| **BEAT-AD**<br>**2016** | 4 | 75.0 | 78.1 (8.0) | 10-20 inclusive | Probable AD<br>NINCDS/ADRDA |
| **van Dyck**<br>**2019** | 80 | 38.8 | >55 | 18-26 inclusive | Probable AD NIAAA |
| **Frolich**<br>**2019** | 43 | 65.1 | 72.2 (6.5) | $\geq 24$ | "Prodromal AD" |
| **REVERSE-SD**<br>**2021** | 83 | 51.8 | 72.6 (NA) | 20-28 inclusive | "Mild AD" |
| **Trailblazer-ALZ**<br>**2021** | 126 | 51.6 | 75.4 (5.4) | 20-28 inclusive | "Prodromal AD" |
| **Wang**<br>**2021** | 22 | 50.0 | 71.3 (6.7) | 16-26 inclusive | Probable AD NIAAA |

Several progression measures were excluded due to being used in two or fewer studies. These were APCC, ECOG, EQ-5D, iADL, RBANS, ZCI-AD, ADCS-ADLPI and ADCOMS.

In the preliminary analysis an assessment was made of the heterogeneity of variance across studies for each cognitive outcome measure. The I2 statistic and Q-statistic were used in this analysis. This showed high heterogeneity of variance across studies for nearly all the selected measures. For example, CDR-SB (I2 91.62% and Q-statistic 202.44 p<.0001) and MMSE (I^2^ 86.69% and Q-statistic 112.69 p<.0001). Only one measure appeared to have low heterogeneity of variance across studies (six studies), ADCS-ADL (I^2^ 0% and Q-statistic 3.22 p 0.665).

Given the high heterogeneity between the studies, a random effects model [17] was utilised for the main meta-regression to account for this. This model assumes that different studies estimate different effects but that these are related with a resultant adjustment to study weighting according to the extent of heterogeneity, or variation, among the effects [10]. The random effects model was further adjusted by using inverse variance to assign weights to each study.

The results of the meta-analysis for individual progression measures are detailed in **Figure 2**. In each panel, a group of measures is meta-analysed, and modifiers corrected for (thus meta-regressed) when possible. ADCS-ADL-MCI showed no variation in AD stage across the studies utilising the tool, and thus only percentage of females and mean age were regressed. Neither QOL-AD nor ADAS-COG 14 were used in sufficient studies to enable correcting for all three variables. Upon assessment, it was decided that the mean age at baseline and percentage of females at baseline for the studies did not differ significantly, and therefore only AD stage was regressed. The remaining nine measures underwent correction for all three modifiers. The result of the meta-regression is further displayed below each progression measure group to allow for more detail to be visualised.

**Figure 2:**
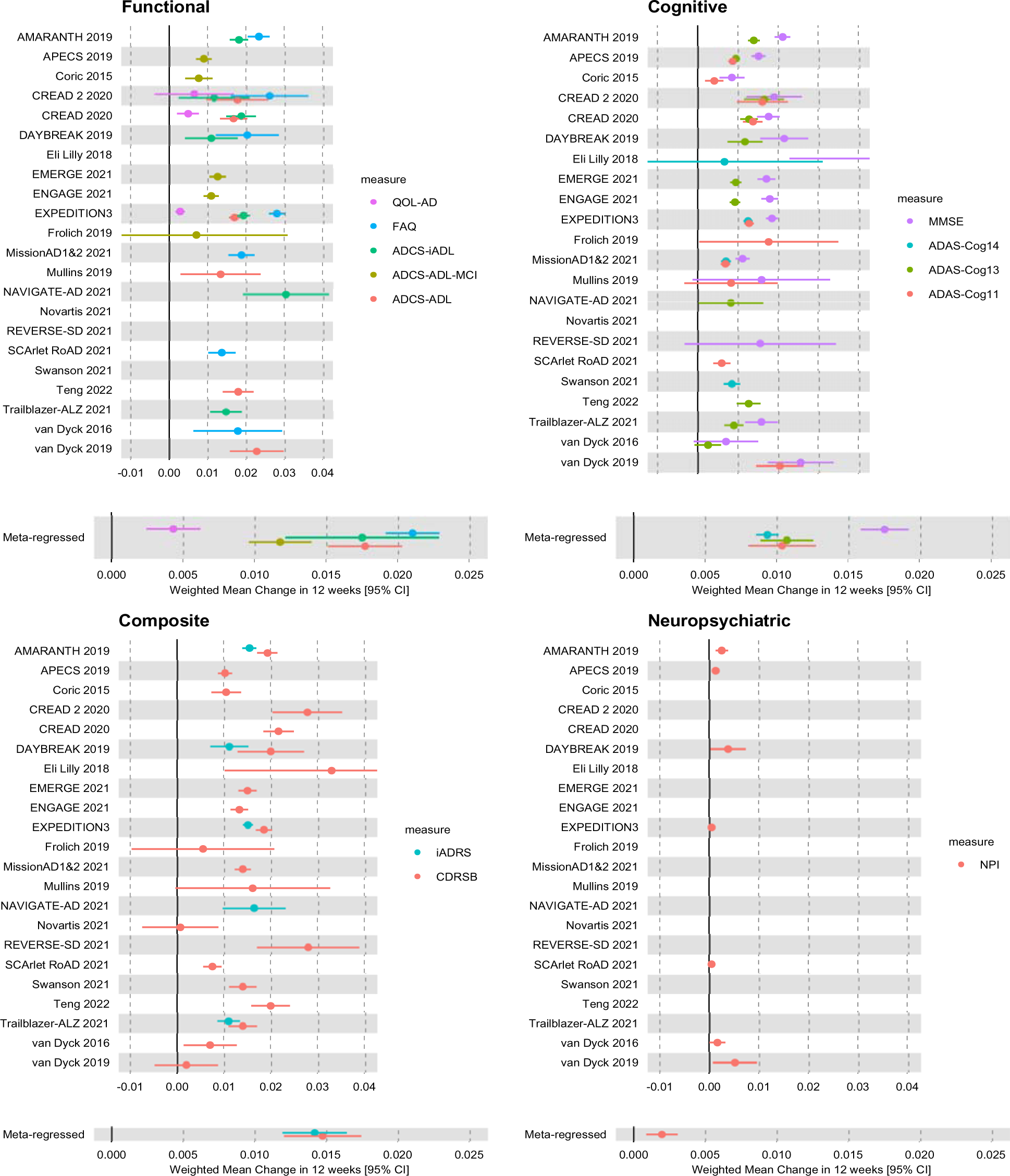
Forest plots of meta regression for functional, cognitive, composite and neuropsychiatric measures from 25 RCTs. Each panel depicts one group of progression measures with their respective meta regression results displayed under the panel for clarity.

It can be seen from the meta regressions that functional measures outperform cognitive and composite measures whilst neuropsychiatric batteries barely detect any change in the short period of time assessed.

Additionally, **Table 2** lists outcome measures by weighted mean change over 12 weeks. A comparison of raw weighted mean change and the same with modifiers accounted for shows the necessity of correcting for age, gender, and AD stage.

**Table 2:**
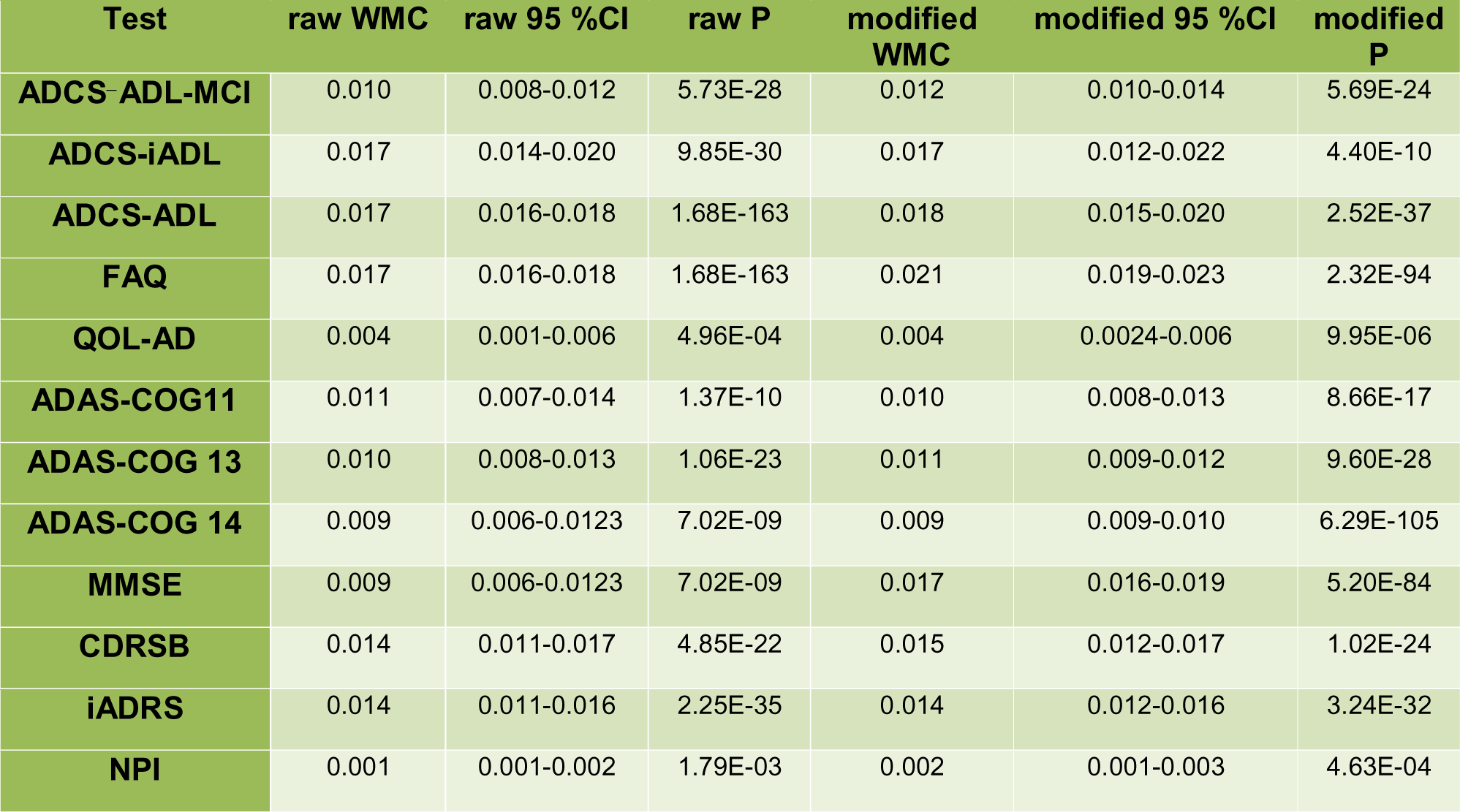
Results of the meta analysis as well as meta regression for each progression measure. WMC=weighted mean change. P-values calculated from Z-scores.

## Discussion

In this study we performed meta-analyses of cognitive, functional, composite, and neuropsychiatric tests used in RCTs that met our inclusion criteria. Only the placebo arm of the RCTs with amyloid positive subjects were included in this analysis. Whilst all disease stages were covered in this meta-analysis, most of the included studies targeted early and prodromal phases of AD. To our knowledge, no similar systematic review or meta-analysis exists at this scale. Twenty-five RCTs were included in this study spanning a publication period from 2015 to 2022.

Overall, measures that assess subject ADLs wholly or as part of a composite score, offer the best performance in detecting progression in subjects. The FAQ, an 18 point functional measure, had the largest weighted mean change over the 12-week time period (0.21, 95% CI 0.019 - 0.023). FAQ has been previously shown to be reliable, valid, and helpful in differentiating between normal controls, MCI and mild dementia [18].

The FAQ is a subjective measure with potential for greater variability in rating. It is administered to informants, and it has been demonstrated that informants cohabiting with the subject, and with higher educational attainment, tend to score the subject with greater impairment, particularly where the subject has MCI [19].

The performance of the FAQ would appear to be at odds with the modest performance of another purely functional measure in our analysis, the QoL-AD (0.04, 95% CI 0.0024-0.006). The QoL-AD is administered to the subject and contains a mix of neuropsychiatric items, social items, relationship items and functional items. This is unlike the FAQ which has a greater focus on functional performance and is administered to informants. Potentially, the performance of the QoL-AD is attenuated by the inclusion of some of these items (*e.g.,* neuropsychiatric symptoms), uncommon in early disease stages. This would also explain the poor performance of the Neuropsychiatric Inventory (NPI) demonstrated in this analysis. It was previously assumed that measures with a larger scale are more likely to detect small amounts of change, however, in the case of both the NPI and iADRS we see this to be invalid. This highlights the need for clinical measures to be geared towards the symptoms experienced at various stages of disease and that factors such as the study population play a key role in influencing the validity of an instrument for measuring progression in AD.

The ADCS-ADL and its instrumental subscale, the ADCS-iADL, performed similarly (Table 2). They appear to be good measures for detecting progression. Notably, they outperformed the ADCS-MCI modification, specifically calibrated to pick up progression more readily in MCI subjects. Both measures are functional scales, and this again highlights the prominence of such scales in our analysis. Our work supports the notion that functional difficulties are present in early AD and MCI, as other work has shown [20], and that they can be identified with standard outcome measures like the FAQ.

The ADAS-COG, and the various versions of it, are reported to be the most commonly used primary outcome instrument in AD clinical trials [21]. However, they have been criticised for having low sensitivity to detect change and significant ceiling effects in mildly, or minimally, impaired cohorts [22]. This has also been highlighted in pre-dementia cohorts [23] and was consistent with our findings above. Most notably, ADAS-COG 14 appears to be least sensitive to change and is the third least sensitive measure of progression examined in this study. Other measures, such as the CDRSB, were said to be better in these patient groups, with one head-to-head analysis of ADAS-COG and CDRSB showing the latter to be better at detecting treatment differences in AD clinical trials [24]. Unlike our study, this work did not analyse RCTs where Aβ*+* status was an inclusion criterion and did not correct for possible modifiers of effect. These are significant limitations. In our findings all three versions of the ADAS-COG displayed inferior ability to measure progression than the CDRSB. The CDRSB contains three functional measures alongside three cognitive measures and although it has been said that the functional measures are not likely to be affected in early stages of AD [25] in our findings this was clearly not the case.

To enhance the ADAS-COG for populations with MCI and early AD, Raghaven and colleagues [26] derived composite measures using the ADAS-COG. They found that the power to detect change could be improved by adding functional items consistent with the superior performance of almost all functional measures in our study. One of the principal measures used to augment the ADAS-COG was the FAQ. This would appear to be a good choice as evidenced by our work.

It has been noted that composite scores, including the iADRS, may outperform their components when determining effect sizes of interventions [27]. In our analysis the iADRS outperformed one component, the ADAS-COG 14, but not the other, the ADCS-iADL. There has been criticism of the proliferation of composite scores in AD trials and that many are not validated before use. Specifically, criticism has focused on lack of available data on performance of the individual components of composite scores [28]. This review goes some way to addressing this issue.

Of the purely cognitive measures, the 30 point Mini-Mental State Examination (MMSE) performed better than expected given the previously stated ceiling effects. The MMSE is well known to have ceiling effects [29] and is affected by education levels [30]. It is likely that the pooled placebo subjects in our study have higher levels of education as has been shown across previous pooled AD trial data [31]. Ceiling effects may be reinforced as a result; however, educational attainment was not readily available across all RCTs to allow for correction of such effects.

A major strength of this work is that it offers a naturalistic view of how established outcome measures assess progression in AD with confirmed AD pathology. This gives an indication of what rate of change in these measures can be expected.

The results should be considered in the context of potential limitations. Whilst every effort was made to find trials for inclusion, there may be studies and data sets which have not been included. The quality of studies included has been rigorously assessed using the Cochrane Risk of Bias tool and this has shown some to be at high risk of potential bias. The inclusion of grey literature is a strength of the work.

The decision to look at placebo patients potentially leads to placebo effects in the pooled cohort. The placebo effect in such patients has already been described with regard to neuropsychiatric symptoms [32]. Some patients may have been taking cholinesterase inhibitors, although with most trials including early-stage patients, this is unlikely to have been a significant proportion.

The choice of 12 weeks as the period over which to assess measurement of progression was a pragmatic decision. This was chosen to maximise the incorporation of more RCTs in the analysis. Our results, when considered over 12 months, are in line with reported minimally clinically important differences (MCID) in the main outcome measures [33].

Additional moderator analyses such as educational attainment or inter-rater variability may be relevant, but were not feasible for every meta-analysis performed, due to lack of information across all studies and degrees of heterogeneity in trial methodology. We believe that those chosen were the most feasible with the given data and also likely to have modifying effects on estimates.

In conclusion, progression measures which rely on purely cognitive domains are not optimal for use in AD trials. Measures should include both cognitive and functional components. Indeed, functional measures appear to be more important. It is not unreasonable to decide to use individual cognitive and functional measures. Most trials included in this analysis have used a range of measures.

The performance of NPI and QoL-AD tests indicate that measures which contain neuropsychiatric items are unlikely to show detectable change over a short period of time, particularly in earlier and presymptomatic AD states which were the predominant cohort in this study, and thus should not be routinely used. The ADCS-ADL, and its instrumental component ADCS-iADL, appear to be functional measures which perform well in detecting change in Aβ*+* enriched subjects in early AD states. As composite scores, the iADRS, which combines the ADAS-COG 14 and the ADCS-iADL, and the CDRSB show good promise for future trial design.

## Data Availability

All data is publicly available and summarised in table 1. Code and raw data will be available from https://github.com/MaryamShoai/hardy-lab-statistical-genetics.github.io in due course.

## Acknowledgement

We are deeply grateful to all those who played a role in the success of this project. In particular we would like to thank Ms Sonia Ruiz Garcia for her invaluable input and support. Her insights and expertise were instrumental in adapting the analysis code for validation by others.

## Appendix 1 Abbreviation of measurements obtained for meta regression

| Abbreviation | Measure Name |
| --- | --- |
| **ADAS-COG 11** | Alzheimer's Disease Assessment Scale–Cognitive Subscale -11 |
| **ADAS-COG 13** | Alzheimer's Disease Assessment Scale–Cognitive Subscale -13 |
| **ADAS-COG 14** | Alzheimer's Disease Assessment Scale–Cognitive Subscale -14 |
| **ADCOMS** | Alzheimer's Disease Composite Score |
| **ADCS ADL-PI** | Alzheimer's Disease Cooperative Study ADL prevention instrument |
| **ADCS-ADL-MCI** | Alzheimer's Disease Cooperative Study – Activities of Daily Living for Mild Cognitive Impairment (ADCS-MCI-ADL) |
| **ADCS-ADL** | Alzheimer's Disease Cooperative Study-Activities of Daily Living inventory |
| **ADCS-iADL** | Alzheimer's Disease Cooperative Study-Activities of Daily Living inventory, instrumental items |
| **APCC** | Alzheimer's Prevention Initiative Composite |
| **CDRSB** | Clinical Dementia Rating Scale Sum of Boxes |
| **ECOG** | Measurement of Everyday Cognition |
| **EQ-5D** | EuroQol 5-dimension questionnaire |
| **FAQ** | Functional Activities Questionnaire |
| **iADL** | Instrumental Activity of Daily Living |
| **iADRS** | Integrated Alzheimer's Disease Rating Scale |
| **MMSE** | Mini-Mental State Examination |
| **NPI** | Neuropsychiatric Inventory |
| **NTB** | neuropsychological test battery |
| **QOL-AD** | Quality of Life in Alzheimer's Disease |
| **RBANS** | Repeatable Battery for the Assessment of Neuropsychological Status |
| **ZCI-AD** | Zarit Caregiver Interview for Alzheimer's Disease |

## Appendix 2 Scoping searches

### Alzheimer’s disease Clinical trials with Patient Amyloid Status documented (By PET or CSF)

**Ovid MEDLINE(R) <1946 to April 8^th^ 2022>**

1 alzheimer*.ti. or exp *alzheimer disease/

2 (cerebrospinal fluid or CSF).ti.

3 Positron-Emission Tomography/

4 exp *amyloid/

5 2 or 3 or 4

6 1 and 5

7 (prognos* or progression).tw.

8 exp prognosis/

9 exp disease progression/

10 exp epidemiologic studies/

11 7 or 8 or 9 or 10

12 6 and 11

13 controlled trial.tw.

14 (cohort or prospective or retrospective or follow-up or longitudinal).tw.

15 cross-sectional.tw. or cross-sectional/

16 Pragmatic Clinical Trial/ or Clinical Trial, Phase II/ or Controlled Clinical Trial/ or Clinical Trial, Phase I/ or Randomized Controlled Trial/ or Clinical Trial/ or Clinical Trial, Phase III/ or Clinical Trial, Phase IV/

17 (drug or therapeutic or medication).mp. [mp=title, abstract, original title, name of substance word, subject heading word, floating sub-heading word, keyword heading word, organism supplementary concept word, protocol supplementary concept word, rare disease supplementary concept word, unique identifier, synonyms]

18 13 or 14 or 15 or 16 or 17

19 18 not 15

20 12 and 19

21 (mouse or mice or murine or rat or rats or animal or rodent or monkey or primate or drosophila).ti.

22 20 not 21

23 limit 22 to yr=“2005 -Current”

24 limit 23 to english language

**Total retrieved= 1687**

### Alzheimer’s disease Clinical trials with Patient Amyloid Status documented (By PET or CSF)-version 2

**Ovid Embase <1974 to April 8^th^ 2022>**

1 alzheimer*.ti. or exp *alzheimer disease/

2 (cerebrospinal fluid or CSF).ti.

3 Positron-Emission Tomography/

4 exp *amyloid/

5 2 or 3 or 4

6 1 and 5

7 (prognos* or progression).tw.

8 exp prognosis/

9 exp disease progression/

10 exp epidemiologic studies/

11 7 or 8 or 9 or 10

12 6 and 11

13 controlled trial.tw.

14 (cohort or prospective or retrospective or follow-up or longitudinal).tw.

15 cross-sectional.tw. or cross-sectional/

16 Pragmatic Clinical Trial/ or Clinical Trial, Phase II/ or Controlled Clinical Trial/ or Clinical Trial, Phase I/ or Randomized Controlled Trial/ or Clinical Trial/ or Clinical Trial, Phase III/ or Clinical Trial, Phase IV/

17 (drug or therapeutic or medication).mp. [mp=title, abstract, original title, name of substance word, subject heading word, floating sub-heading word, keyword heading word, organism supplementary concept word, protocol supplementary concept word, rare disease supplementary concept word, unique identifier, synonyms]

18 13 or 14 or 15 or 16 or 17

19 18 not 15

20 12 and 19

21 (mouse or mice or murine or rats or animal or rodent or monkey or primate or drosophila).ti.

22 20 not 21

23 limit 22 to yr=“2005 -Current”

24 limit 23 to english language

**Total retrieved= 1495**

**Deduplicated= 2744**

